# Side effects of Covishield vaccine among frontline healthcare workers of a tertiary health care center

**DOI:** 10.1101/2022.06.09.22276129

**Authors:** Durga Dhungana, Yukta Narayan Regmi, Deependra Shrestha, Krishna Thapa, Chandra Bahadur Pun, Tirthalal Upadhayaya, Gopi Hirachan

## Abstract

**Objectives:** COVID disease started in the late 2019 and within a short time became a pandemic disease. With the increasing morbidity and mortality all over the world and the therapeutics not doing wonders, scientists were in the attempt to develop vaccines as a mitigating measure. With continuous efforts and developments, different vaccines were developed and rolled out gradually in different countries. Concerns were notable for occurrence of side effects. Hence this study was done to assess the side effects following Covishield vaccination in Nepal at the initial stage.

**Methods:** This was a cross-sectional study done via snowball sampling method among healthcare workers at a tertiary medical college hospital in Pokhara, Nepal after obtaining ethical consent from the institutional review committee of the concerned hospital. The proforma was sent via online means through different social media platforms and also printed forms were also given to the respondents. A total of 139 respondents were obtained after removing duplications. The data were entered into SPSS and analyzed using descriptive and inferential statistics. P-value ≤ 0.05 was considered statistically significant.

**Results:** Majority (64.7%) were female healthcare workers. More than half (52.3%) used pre-medication in an attempt to avoid the side effects of vaccine. Most (90.6%) reported at least one side effect-local or systemic to the first dose and approximately three-quarter (74.3%) reported side effect to the second dose. Common side effects were pain at injection site, muscle pain, headache, fatigue and weakness. Most of the side effects were higher with the first dose as compared to the second dose.

**Conclusion:** Side effects are common with Covishield vaccination, significantly more with the first dose as compared to the second dose. Female gender, younger age and past covid infection were associated with slightly more occurrence of side effects; however were not found to be statistically significant.

## Background

Coronavirus disease 2019 (COVID-19) started in late 2019 and caused a pandemic around the world. Globally by 3 May 2022, there have been 511,965,711 confirmed cases of COVID-19, including 6,240,619 deaths, reported to World Health Organization (WHO). As of 30 April 2022, a total of 11,532,661,625 vaccine doses have been administered^1^. Nepal is also affected due to COVID-19 with significant morbidity and mortality till date. The first 2019 novel coronavirus case in Nepal was on January 2020. As per the daily update provided by Ministry of Health and Population, Nepal, there were 1119277 confirmed cases of COVID-19 with 11,952 deaths as of 22^nd^ May, 2022^2^. With the increasing cases and mortality all over the world, there was dire need of some intervention to halt the effect. With different therapies introduced but none as major success, the introduction of vaccines became a major boom in the fight for covid.

Different vaccines have been made to control COVID-19. The ones currently approved by WHO include Pfizer, AstraZeneca, Covishield, Janssen, Moderna, Sinovac, Sinopharm, Covaxin and various other vaccines are ongoing assessment. As part of initiation of global vaccination program, a coronavirus vaccine was first administered to 90-year-old Margaret Keenan in the UK on December 8, 2020.^3^ Nepal launched the COVID-19 vaccination campaign on 27 January 2021 vaccinating the frontline healthcare workers with Covishield vaccine at the first phase and gradually expanding the vaccination campaign till date^4^. Second dose of the Covishield was provided to the healthcare workers on the end of April^5^. As of May 22, 95.8% of those above 12 years of age have been inoculated with at least 1 dose and 84.7% with full dose.^2^ Various studies have been published in relation to the occurrence of the side effects in internationally.^6-13^ Datas related to side effects have been minimal in context of Nepal.^14-19^ Hence this research is an attempt to address the side effects of Covishield vaccine in healthcare workers working at a tertiary health care center in Nepal.

## Methodology

A cross-sectional study was conducted among the healthcare workers at a tertiary care hospital in Pokhara, Nepal from July 1^st^, 2021 to July 15^th^, 2021 after obtaining ethical approval from the Institutional Review Board of concerned teaching hospital. Assuming 50 percent of the vaccinated healthcare workers will develop adverse effect related to vaccination with 95% confidence and a margin of error of 10%, the sample size calculated using Cochran’s formula:

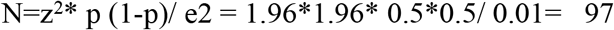

Considering 10 percent as non-response rate, the minimum required sample was 107.

Data was collected via online means via google form and also via printed form of the performa. Those healthcare workers included in the inclusion criteria were contacted via different social platforms like messenger, facebook, Whatsapp and viber for filling the form. Snowball sampling technique was be used. The primary respondents of the study were identified as per the familiarity with the researcher and those respondents were further requested to forward the link of the questionnaire to others who they know and have got two doses of Covishield vaccination.

The semi-structured questionnaire performa containing the sociodemographic details along with multiple options selection questions was prepared based on the extensive literature review and peer review. Open-ended questions of medications taken are included and remaining questions were close-ended.

The form consisted of informed consent at first. If respondent answered no, then no further questions were asked. If respondent answered yes, then it would lead to the other parts of the form. Hence the study questions were followed after obtaining informed consent only. This included demographic characteristics, history of comorbidity, history of past covid infection, use of premeditations prior vaccination and work area involved. This was followed by multiple choice options related questions of the local side effects that occurred within the first 24 hrs and their persistence up to 7 days and beyond 7 days. This was followed by multiple choice options related questions of the systemic side effects that occurred within the first 24 hrs and their persistence up to 7 days and beyond 7 days.

Total of 160 responses were collected. After obtaining the responses, the data were entered into Microsoft Excel and checked for duplications and incomplete responses. After omitting duplications and incomplete responses, 139 responses were finally analysed in SPSS 21. The demographic characteristics were expressed in terms of percentages. For comparison of side effects between different categorical variables like gender, history of covid infection, dosage of Covishield vaccine, chi-square test was used. A p-value ≤ 0.05 was considered statistically significant.

### Principal Findings

#### 1. Sociodemographic characteristics

Majority of the respondents were below 40 years of age. Approximately ten percent of the respondents used medicines in an attempt to avoid the side effects. Use of medications for symptom relief was seen in nearly half of the individuals after receiving the first dose of Covishield vaccine. Majority of the respondents developed side effects following the first dose whereas a quarter of those receiving the second dose didn’t develop side efffect. (Table 1)

**Table 1.**
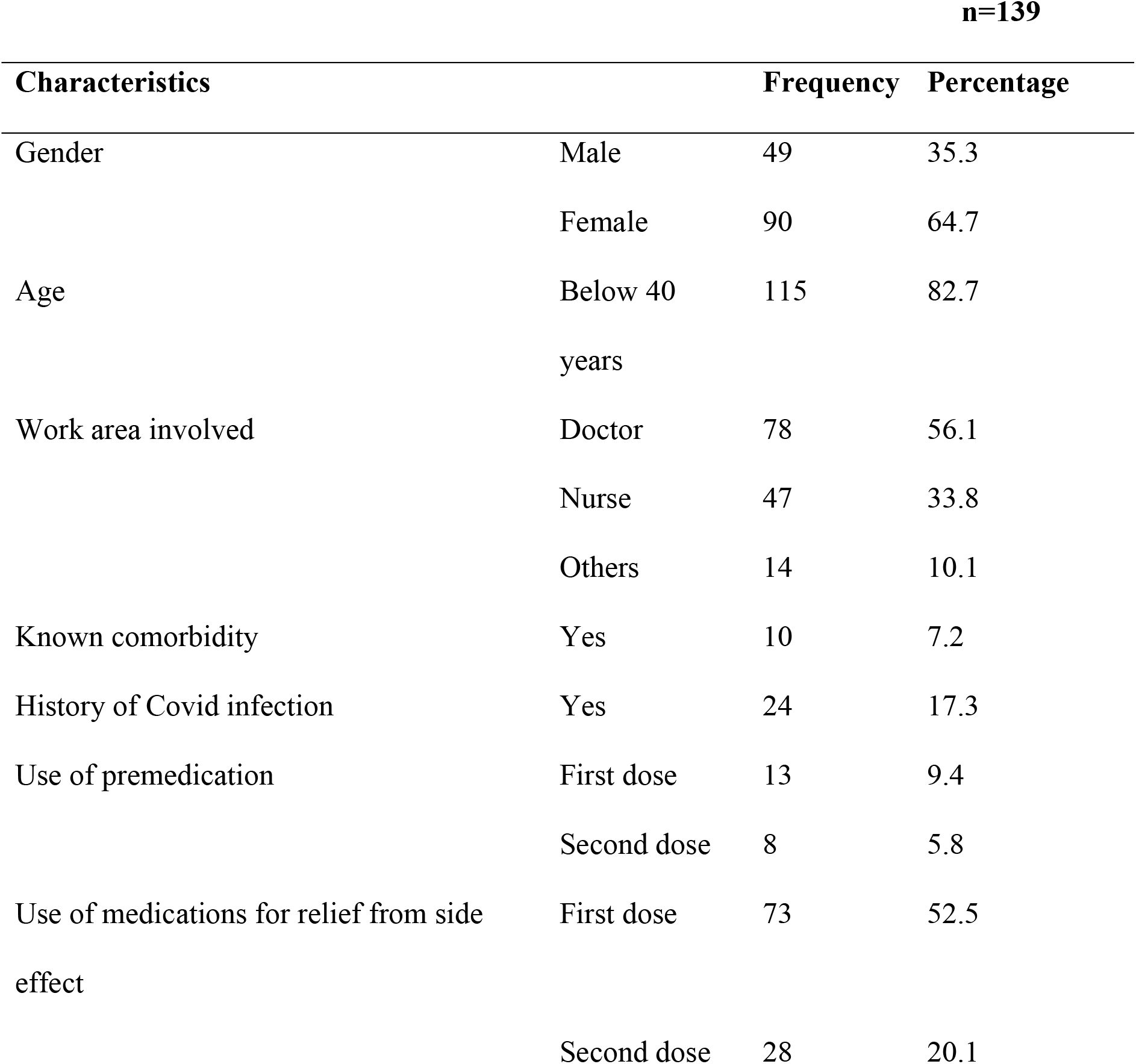
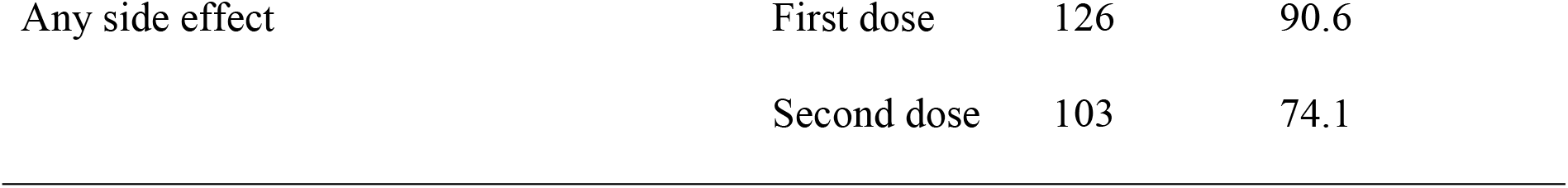
Basic characteristics of the participants

#### 2. Local side effects

Pain at the injection site was the most common local side effect reported. Most of the local side effects except itching were more common after the first dose received. (Table 2)

**Table 2.** Frequency of local side effects (1st and 2nd Covishield dose)

| Characteristics | First dose | Second dose | P-value |
| --- | --- | --- | --- |
| Number (percentage) |  |  |  |
| Local effects | 105 (75.5) | 86 (61.8) | **.02** |
| No side effects | 34 (24.5) | 53 (38.2) |  |
| Pain | 101 (72.6) | 84 (60.4) | **.04** |
| Swelling | 11 (0.08) | 8 (0.06) | .64 |
| Redness | 8 (0.06) | 5 (0.03) | .57 |
| Itch | 5 (0.03) | 11 (0.08) | .2 |

#### 3. Systemic side effects

Systemic side effects were also more common after the first dose of Covishield vaccine. Muscle pain was most common reported symptom. (Table 3)

**Table 3.** Frequency of systemic side effects (1^st^ and 2^nd^ Covishield dose)

| Characteristics | First dose | Second dose | P-value |
| --- | --- | --- | --- |
|  | Number (percentage) |  |  |
| Systemic effects | 121(87) | 77 (55.4) | $\leq 0.001$ |
| No side effects | 18 (13) | 62 (44.6) |  |
| Muscle pain | 80 (57.5) | 45 (32.4) | $\leq 0.001$ |
| Fatigue | 66 (47.5) | 23 (16.5) | $\leq 0.001$ |
| Headache | 65 (46.7) | 33 (23.7) | $\leq 0.001$ |
| Weakness | 59 (42.4) | 23 (16.5) | $\leq 0.001$ |
| Fever | 50 (35.9) | 3 (2.1) | $\leq 0.001$ |
| Dizziness | 33 (23.7) | 15 (10.8) | .007 |
| Joint pain | 21 (15.1) | 6 (4.3) | .004 |
| Nausea | 21 (15.1) | 11 (7.9) | .09 |

#### 4. Sociodemographic variables comparison with dose of vaccine

On comparison of side effects both local and systemic of different Covishield doses with different sociodemographic variables, no factors were found to be significantly associated except for systemic side effects after first dose in those aged below 40 years. (Table 4)

**Table 4.**
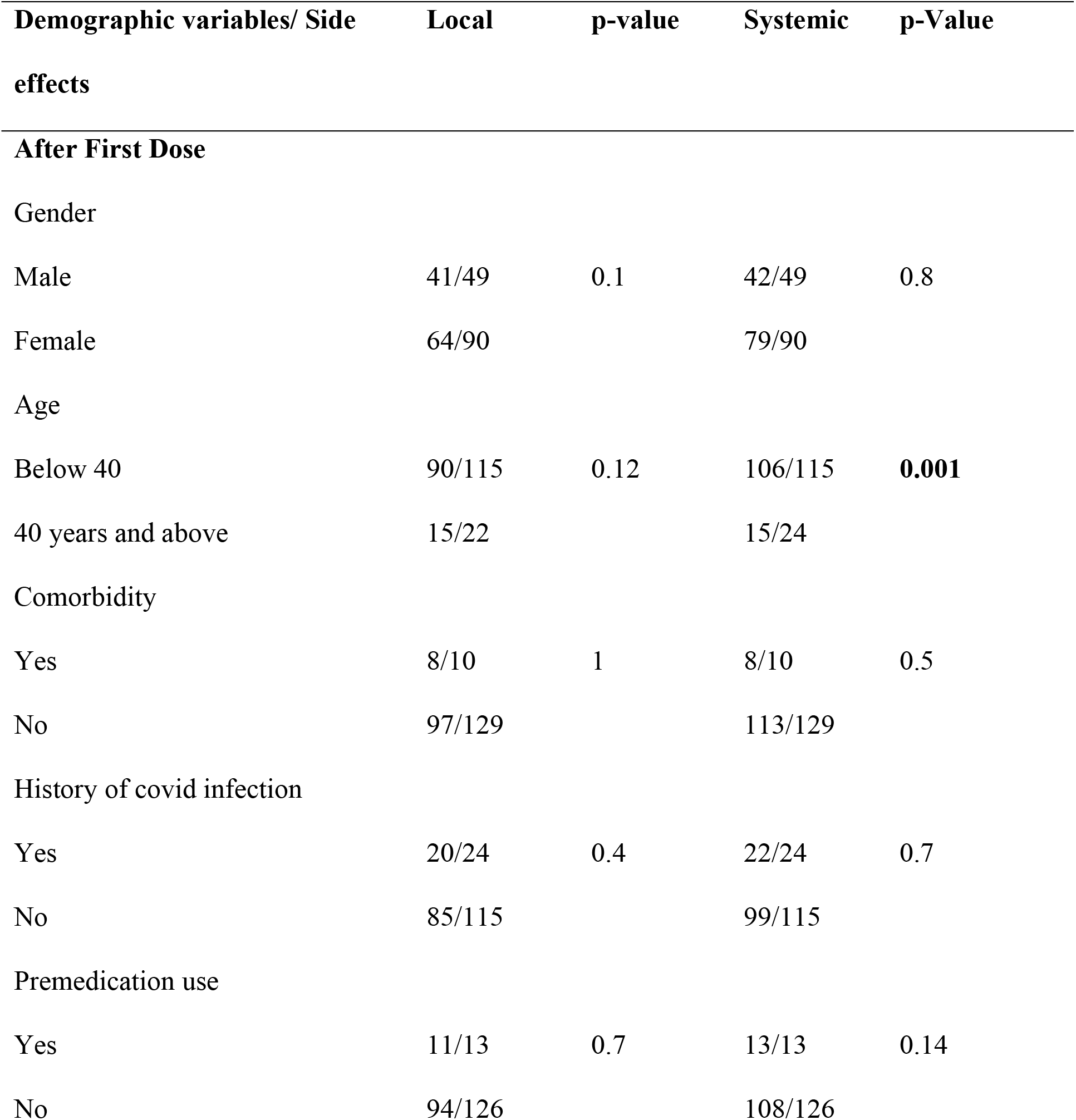
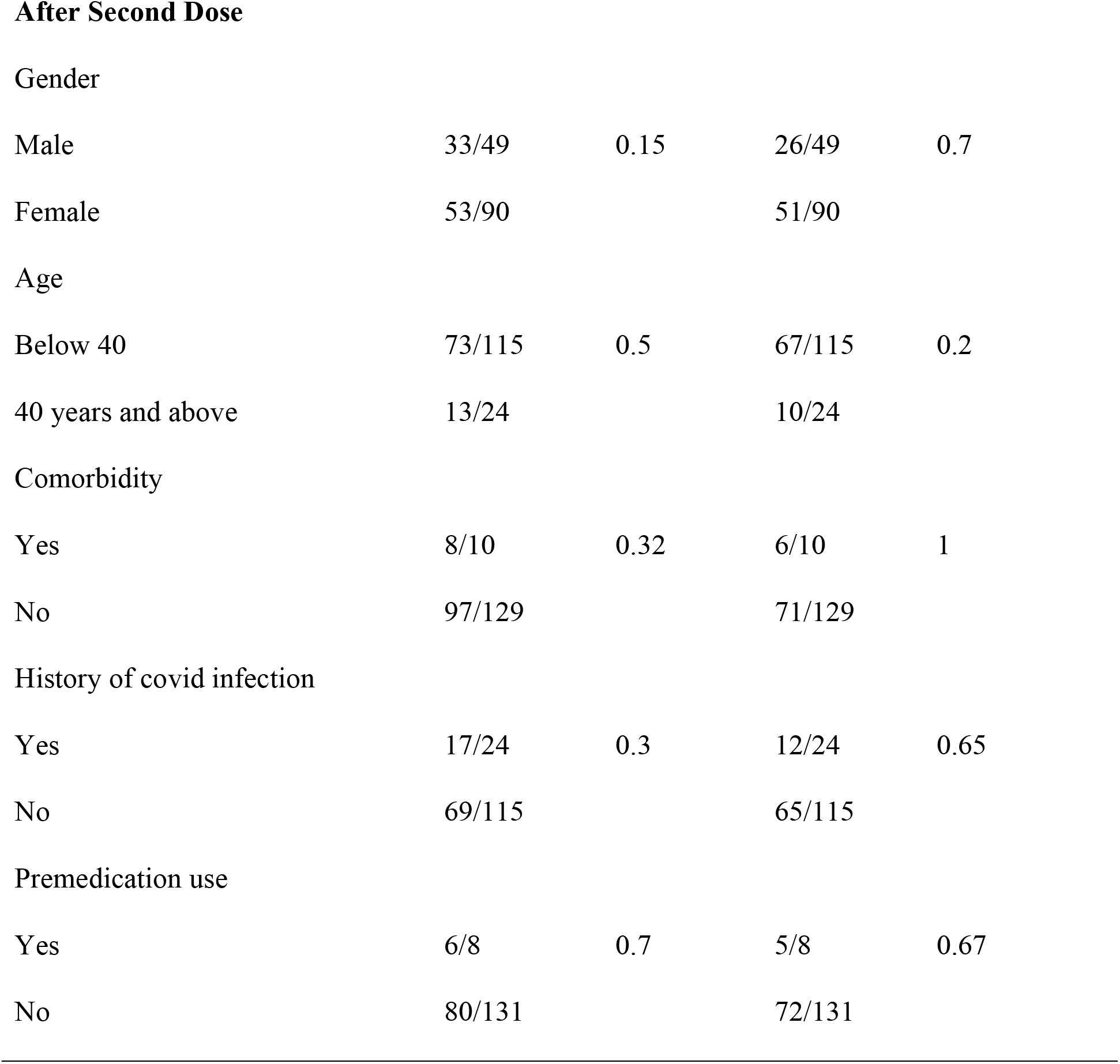
Cross tabulation of side effects after doses 1 and 2 with demographic variables

## Discussion

Among the respondents, female were the majority one which is also similar to other studies done among healthcare workers in different countries. ^6, 8, 12-14, 20^ Females are found to be more responding than males.

Common side effects being reported were pain at injection site, muscle pain, fatigue, headache and weakness. This was similar to side effects reported to same or other vaccines in other studies. ^7-13, 21, 22^ However the occurrence of side effects was less in healthcare workers in Turkey who received CoronaVac vaccine and also lower in those receiving Sputnik vaccine.^7, 20^ Similar to the Pfizer-BioNTech vaccination of healthcare workers at Malta’s state hospital, local side effects were common with the first dose as compared to the second dose. However the systemic side effects were common with the first dose of Covishield vaccine unlike Pfizer vaccine.^8^ Similar to the side effects reported by healthcare workers in Iran who received first and second dose of Sputnik vaccine and Egyptian population receiving first and second dose of AstraZeneca vaccine, the frequency of side effects like injection site pain, fever, headache, body pain, fatigue were more among those receiving the first dose as compared to second dose and was statistically significant (p<0.05).^7, 9^ This is also in congruence with the study done among Jordanian healthcare workers who reported more occurrence of systemic side effects with first dose of AstraZeneca vaccine as compared to the second dose.^6^

Similar to the study done in Patan of Nepal, among those receiving first dose of Covishield in the rollout vaccine campaign, more than half (52.5%) used some medications for symptom relief for the side effect. ^14^ Those with prior history of covid had higher frequency of side effects both local and systemic, however was statistically insignificant and this was similar to the study done in Iran for the Covishield group and also in Patan of Nepal. ^14, 22^ This finding is in contrast to the study done in a university tertiary hospital in France. Possible reason may be due to the difference in the type of vaccine used and difference in the study population.^21^ Similar to other studies, there was no significant difference in occurrence of side effects between males and females. ^14, 22^ Nonetheless, female healthcare workers in Iran receiving Sputnik vaccine reported statistically significant higher occurrence of side effects as compare to the male groups.^7^ Possible reason may be due to the difference in the study population, timing of the study and the difference in the type of vaccine. On analysis it was found that those aged below 40 years reported more side effects than the group aged above 40 years. However it was statistically significant only for the systemic effects following the first dose. These findings were similar to other studies which showed higher prevalence of side effects in those below 40 years of age. ^7, 8, 12, 13, 22^ However German healthcare workers of older age group reported more systemic side effects as compared to younger age group among the viral-vector based COVID-19 vaccine. ^11^ Presence of comorbidities didn’t affect the occurrence of side effects and was similar to the finding among Turkish healthcare workers. ^20^

The limitations of this study are it is a single-center study with moderate sampling size. The respondents were asked about the side effects that occurred during the time of vaccination so recall bias can occur. The side effects though given options were self-reported by the respondents hence direct face-to-face questionnaire wasn’t done. This could cause the respondent to report only the symptoms which the patient thought important and worrisome. Also the sampling method was snowball sampling method hence may not be actual representation of the targeted population.

## Conclusions

Side effects are common following Covishield vaccination. Commonest side effects are pain at injection site, malaise, headache and muscle pain. The side effects are more pronounced with the first dose of Covishield vaccine in comparison to the second dose. Past COVID infection, presence of comorbidities, age, use of pre-medication for side effect relief and gender didn’t have a significant difference in the occurrence of side effects.

## Data Availability

The data is made available as supporting file.

## Ethical approval

The study has been approved by the institutional ethical committee of Gandaki Medical College Teaching Hospital and Research Center Private Limited (Ref no: 10/2078/2079) on 30^th^ June, 2021. Subjects gave informed consent to the study.

## Acknowledgement

None

## Data availability

The data that support the findings of this study are available from the corresponding author upon reasonable request.

